# Demographic and health characteristics associated with fish and n-3 fatty acid supplement intake during pregnancy: results from pregnancy cohorts in the ECHO program

**DOI:** 10.1101/2023.11.17.23298695

**Authors:** Emily Oken, Rashelle J. Musci, Matthew Westlake, Kennedy Gachigi, Judy L. Aschner, Kathrine L. Barnes, Theresa M. Bastain, Claudia Buss, Carlos A. Camargo, Jose F. Cordero, Dana Dabelea, Anne L. Dunlop, Akhgar Ghassabian, Alison E. Hipwell, Christine W. Hockett, Margaret R. Karagas, Claudia Lugo-Candelas, Amy E. Margolis, Thomas G. O’Connor, Coral L. Shuster, Jennifer K. Straughen, Kristen Lyall, program collaborators for Environmental influences on Child Health Outcomes

## Abstract

**Objective:** Omega-3 (n-3) fatty acid consumption during pregnancy is recommended for optimal pregnancy outcomes and offspring health. We examined characteristics associated with self-reported fish or omega-3 supplement intake.

**Design:** Pooled pregnancy cohort studies.

**Setting:** Cohorts participating in the Environmental influences on Child Health Outcomes (ECHO) consortium with births from 1999-2020.

**Participants:** A total of 10,800 pregnant people in 23 cohorts with food frequency data on fish consumption; 12,646 from 35 cohorts with information on supplement use.

**Results:** Overall, 24.6% reported consuming fish never or less than once per month, 40.1% less than once a week, 22.1% 1-2 times per week, and 13.2% more than twice per week. The relative risk (RR) of ever (vs. never) consuming fish was higher in participants who were older (1.14, 95% CI: 1.10, 1.18 for 35-40 vs. <29 years), were other than non-Hispanic White (1.13, 95% CI: 1.08, 1.18 for non-Hispanic Black; 1.05, 95% CI: 1.01, 1.10 for non-Hispanic Asian; 1.06, 95% CI: 1.02, 1.10 for Hispanic), or used tobacco (1.04, 95% CI: 1.01, 1.08). The RR was lower in those with overweight vs. healthy weight (0.97, 95% CI: 0.95, 1.0). Only 16.2% reported omega-3 supplement use, which was more common among individuals with a higher age and education, a lower BMI, and fish consumption (RR 1.5, 95% CI: 1.23, 1.82 for twice-weekly vs. never).

**Conclusions:** One-quarter of participants in this large nationwide dataset rarely or never consumed fish during pregnancy, and omega-3 supplement use was uncommon, even among those who did not consume fish.

## INTRODUCTION

Omega-3 (n-3) polyunsaturated fatty acids (PUFAs) are essential nutrients. Adequate consumption is vital in pregnancy, as omega-3 PUFAs, in particular long-chain docosahexaenoic acid (DHA), contribute to offspring neurodevelopment and may improve pregnancy outcomes, including risk for preterm birth.(^1^) Fish and other seafood (hereafter “fish”) are the main dietary source of long-chain omega-3 PUFAs. Therefore, current guidance recommends intake of 8-12 ounces (224-336 g, or 2-3 servings) of fish per week during pregnancy,(^2,3^) with the goal of consuming an average of 200 mg/day of DHA.(^4^)

Limited research suggests that few pregnant women consume the recommended amounts of fish or omega-3 PUFAs. The latest United States (US) Food and Drug Administration (FDA) assessment of dietary fish intake was conducted in 2014 and relied on data sources by then already decades old.(^5^) In the 2004 Infant Feeding Practices Study II, the median intake of total fish by pregnant participants, excluding non-fish consumers, was 1.8 ounces/week (about 1 serving per month).(^6^) In the 2013 National Health and Nutrition Examination Survey (NHANES), mean fish intake by pregnant women was 4.6 servings a month.(^7^)

Additionally, studies suggest that fish and omega-3 PUFA intake during pregnancy has been declining over past decades, likely in response to federal advisories about mercury in fish since 2001.(^8,9^) In the NHANES survey, mean DHA intake among women of childbearing age decreased from 56 mg/day in 2003-2004 to 42 mg/day in 2011-2012.(^10^) Intake was highest in women who were non-Hispanic White and had higher education and income levels, similar to demographic patterns in non-population–based cohorts.(^11,12^) Despite the importance of fish consumption during pregnancy, women consume substantially less than men and do not increase intake during pregnancy.(^7,10,13^)

Most experts believe that fish consumption is the optimal way to meet recommendations for adequate omega-3 PUFA intake,(^14^) in part because experimental evidence has not supported offspring developmental benefits of supplementation.(^15,16^) For those who cannot or choose not to eat fish, omega-3 PUFAs supplements are recommended.(^17^) The extent to which pregnant women take omega-3 PUFA supplements is not well described. In addition, it is unclear whether supplement use is more common in those with low fish intake. In the 2003-2012 NHANES surveys, only 9% of pregnant women consumed an omega-3 PUFA supplement, but this information was not presented according to year of pregnancy or by fish intake.(^10^)

We examined data from the National Institutes of Health Environmental influences on Child Health Outcomes (ECHO) program(^18,19^) to address our hypotheses that fish consumption would have declined over the past two decades and that supplement use would be more common among those who did not eat fish.

## METHODS

### Study Design, Sample, and Measures

In October 2022, the ECHO data platform included information on more than 52,000 singleton pregnancies from 69 cohorts across the U.S. and Puerto Rico (Supplementary Figure 1). We included data from 23 cohorts that collected information on fish intake during pregnancy and 35 that collected supplement intake during pregnancy. Within cohorts, pregnancies were included if they had information on either fish intake or supplement use.

### Assessment of Fish and Supplement Intake

We performed a keyword search and form review for food frequency questionnaires that assessed fish intake and any questionnaires that assessed supplement intake. For fish intake, we converted relevant questionnaire items and summed as appropriate to weekly total intake. We then constructed a 4-level categorical variable: 1) never or less than once per month, 2) once per month to less than once per week, 3) one to two times per week, and 4) more than twice per week. Additionally, we created a binary variable of never or less than once per month (which we summarize as “never”) vs. more (“ever”). For supplements, we created a binary variable to indicate any (vs. no) use of supplements with fish oil or omega-3 fatty acids.

### Assessment of Other Characteristics

We captured other variables of interest from harmonized derived tables of maternal self-reported sociodemographic characteristics, including age, race, ethnicity, education, tobacco or nicotine use during pregnancy (Y/N), and pre-pregnancy body mass index (BMI), each of which we categorized as in Table 1.

**Table 1:**
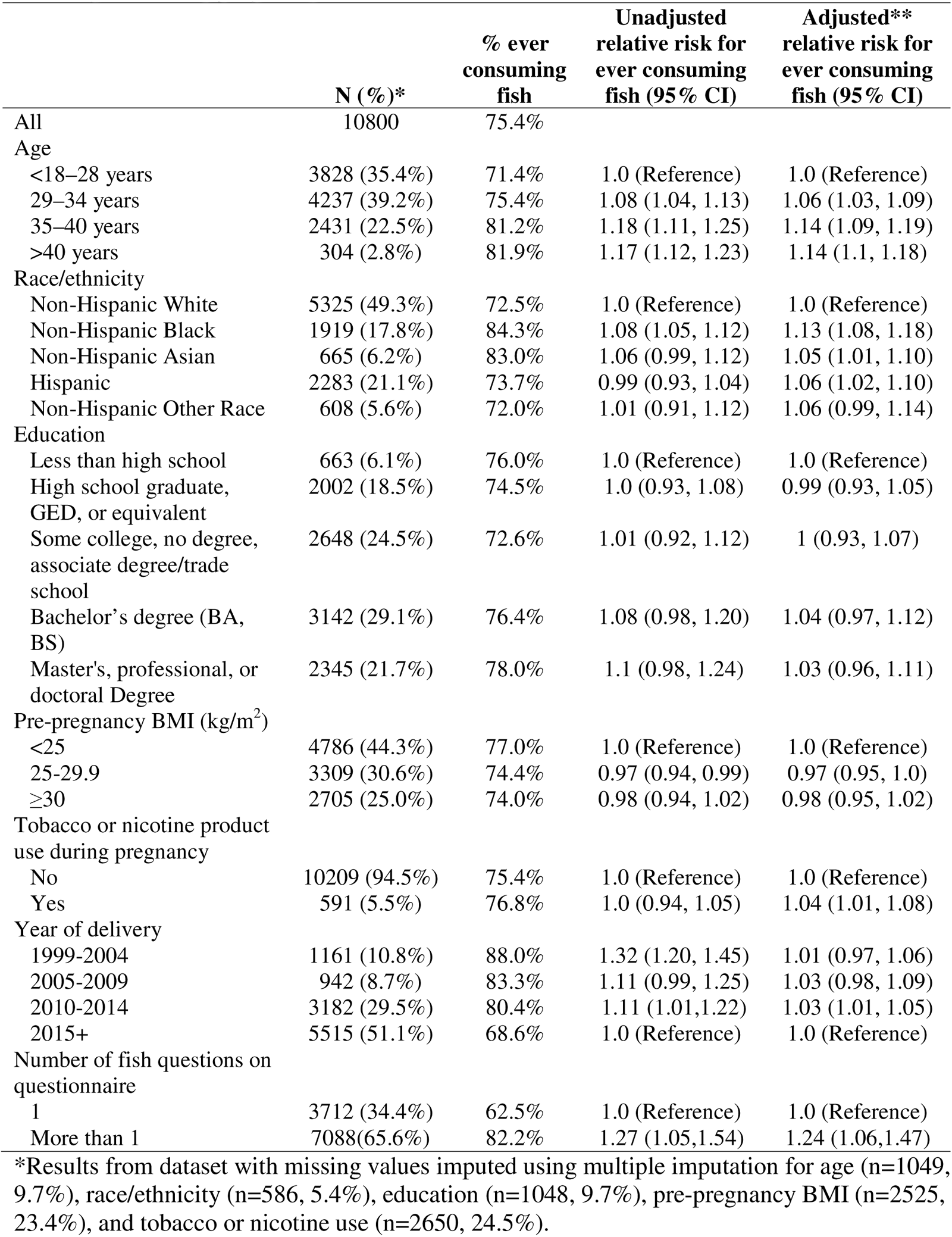
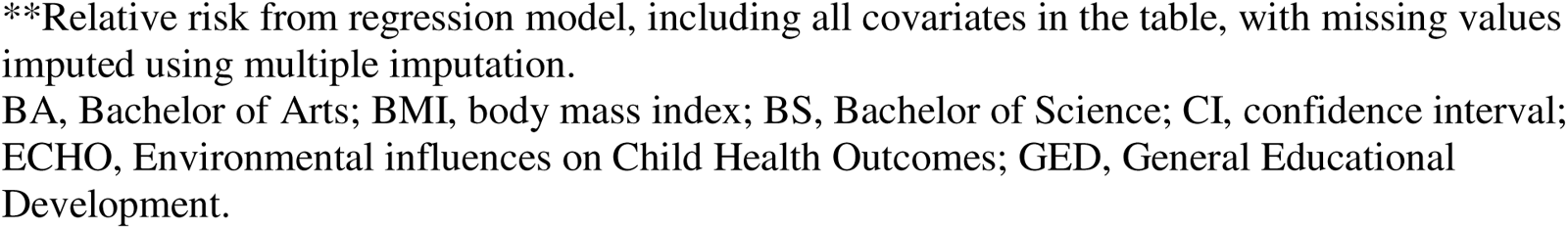
Characteristics of 10,800 ECHO-wide cohort participants with information on fish consumption during pregnancy.

|  | N (%)* | % ever consuming fish | Unadjusted relative risk for ever consuming fish (95% CI) | Adjusted** relative risk for ever consuming fish (95% CI) |
| --- | --- | --- | --- | --- |
| All | 10800 | 75.4% |  |  |
| Age |  |  |  |  |
| <18–28 years | 3828 (35.4%) | 71.4% | 1.0 (Reference) | 1.0 (Reference) |
| 29–34 years | 4237 (39.2%) | 75.4% | 1.08 (1.04, 1.13) | 1.06 (1.03, 1.09) |
| 35–40 years | 2431 (22.5%) | 81.2% | 1.18 (1.11, 1.25) | 1.14 (1.09, 1.19) |
| >40 years | 304 (2.8%) | 81.9% | 1.17 (1.12, 1.23) | 1.14 (1.1, 1.18) |
| Race/ethnicity |  |  |  |  |
| Non-Hispanic White | 5325 (49.3%) | 72.5% | 1.0 (Reference) | 1.0 (Reference) |
| Non-Hispanic Black | 1919 (17.8%) | 84.3% | 1.08 (1.05, 1.12) | 1.13 (1.08, 1.18) |
| Non-Hispanic Asian | 665 (6.2%) | 83.0% | 1.06 (0.99, 1.12) | 1.05 (1.01, 1.10) |
| Hispanic | 2283 (21.1%) | 73.7% | 0.99 (0.93, 1.04) | 1.06 (1.02, 1.10) |
| Non-Hispanic Other Race | 608 (5.6%) | 72.0% | 1.01 (0.91, 1.12) | 1.06 (0.99, 1.14) |
| Education |  |  |  |  |
| Less than high school | 663 (6.1%) | 76.0% | 1.0 (Reference) | 1.0 (Reference) |
| High school graduate, GED, or equivalent | 2002 (18.5%) | 74.5% | 1.0 (0.93, 1.08) | 0.99 (0.93, 1.05) |
| Some college, no degree, associate degree/trade school | 2648 (24.5%) | 72.6% | 1.01 (0.92, 1.12) | 1 (0.93, 1.07) |
| Bachelor's degree (BA, BS) | 3142 (29.1%) | 76.4% | 1.08 (0.98, 1.20) | 1.04 (0.97, 1.12) |
| Master's, professional, or doctoral Degree | 2345 (21.7%) | 78.0% | 1.1 (0.98, 1.24) | 1.03 (0.96, 1.11) |
| Pre-pregnancy BMI (kg/m<sup>2</sup>) |  |  |  |  |
| <25 | 4786 (44.3%) | 77.0% | 1.0 (Reference) | 1.0 (Reference) |
| 25-29.9 | 3309 (30.6%) | 74.4% | 0.97 (0.94, 0.99) | 0.97 (0.95, 1.0) |
| ≥30 | 2705 (25.0%) | 74.0% | 0.98 (0.94, 1.02) | 0.98 (0.95, 1.02) |
| Tobacco or nicotine product use during pregnancy |  |  |  |  |
| No | 10209 (94.5%) | 75.4% | 1.0 (Reference) | 1.0 (Reference) |
| Yes | 591 (5.5%) | 76.8% | 1.0 (0.94, 1.05) | 1.04 (1.01, 1.08) |
| Year of delivery |  |  |  |  |
| 1999-2004 | 1161 (10.8%) | 88.0% | 1.32 (1.20, 1.45) | 1.01 (0.97, 1.06) |
| 2005-2009 | 942 (8.7%) | 83.3% | 1.11 (0.99, 1.25) | 1.03 (0.98, 1.09) |
| 2010-2014 | 3182 (29.5%) | 80.4% | 1.11 (1.01,1.22) | 1.03 (1.01, 1.05) |
| 2015+ | 5515 (51.1%) | 68.6% | 1.0 (Reference) | 1.0 (Reference) |
| Number of fish questions on questionnaire |  |  |  |  |
| 1 | 3712 (34.4%) | 62.5% | 1.0 (Reference) | 1.0 (Reference) |
| More than 1 | 7088(65.6%) | 82.2% | 1.27 (1.05,1.54) | 1.24 (1.06,1.47) |

\*Results from dataset with missing values imputed using multiple imputation for age (n=1049, 9.7%), race/ethnicity (n=586, 5.4%), education (n=1048, 9.7%), pre-pregnancy BMI (n=2525, 23.4%), and tobacco or nicotine use (n=2650, 24.5%).
\*\*Relative risk from regression model, including all covariates in the table, with missing values imputed using multiple imputation.
BA, Bachelor of Arts; BMI, body mass index; BS, Bachelor of Science; CI, confidence interval; ECHO, Environmental influences on Child Health Outcomes; GED, General Educational Development.

### Statistical Analysis

We performed multiple imputation (N=25 imputations) using SAS Proc MI to fill in missing data on covariates of interest. We then performed a log-binomial regression analysis, mutually adjusted for measured demographic characteristics (as in Table 1), smoking status, and pre-pregnancy BMI. We included a random effect for cohort to account for the nested nature of the pooled data and conducted leave-one-cohort-out analyses to confirm that no cohort explained the overall associations. Additionally, we included the number of fish questions asked on the dietary questionnaire (1 or more than 1) given our prior research that found that asking more questions leads to a higher estimate of fish intake.(^20^) Income was not significantly associated with either outcome and including it did not substantially alter any other estimates; therefore, we did not include it in our final models.

## RESULTS

Among 10,800 pregnant people with information on fish consumption, 24.6% reported never consuming fish and 75.4% reported ever consuming fish during pregnancy (Table 1): 40.1% less than 1 serving per week, 22.1% 1-2 servings per week, and 13.2% more than 2 servings per week (Supplemental Table 1). In the multivariable regression analyses with imputed missing covariates (Table 1), the likelihood of ever (vs. never) consuming fish during pregnancy remained higher in people who were older (relative risk [RR] 1.14, 95% CI: 1.10, 1.18 for >40 vs. <29 years), were other than non-Hispanic White (RR 1.13, 95% CI: 1.08, 1.18 for non-Hispanic Black; 1.05, 95% CI: 1.01, 1.10 for non-Hispanic Asian; 1.06, 95% CI: 1.02, 1.10 for Hispanic), had lower BMI (RR 0.97, 95% CI: 0.95, 1.0 for overweight vs. normal BMI), and used tobacco or nicotine products (RR 1.04, 95% CI: 1.01, 1.08). After accounting for demographics and the number of fish questions included on the different dietary questionnaires, no differences in fish intake were observed by year of delivery.

Among 12,646 pregnant people with information on omega-3 PUFA supplement intake, 16.2% reported any supplement use. Supplement use was uncommon before 2005 (less than 0.05%), but not substantially different afterward. In multivariable regression analyses (Table 2), supplement use was more likely at an older age and a higher level of education: those over 40 years of age were about twice as likely to use supplements than those less than 29 years of age (RR 2.01, 95% CI: 1.35, 3.00) and those with a graduate degree were more likely to use supplements than those with less than a high school education (RR 1.71, 95% CI: 1.21, 2.41). Supplement use was less likely among non-Hispanic Black (RR 0.61, 95% CI: 0.48, 0.76) and Hispanic (RR 0.67, 95% CI; 0.55, 0.80) participants compared with non-Hispanic White participants, those who used tobacco or nicotine products (RR 0.81, 95% CI: 0.68, 0.98), and those with a higher BMI (RR 0.79, 95% CI: 0.68, 0.90 for BMI ≥30 vs. <25 kg/m^2^). In contrast to the advice that those who do not consume fish should take an omega-3 fatty acid supplement, supplement use was highest among those with greater fish consumption (Table 2 and Figure 1).

**Figure 1:**
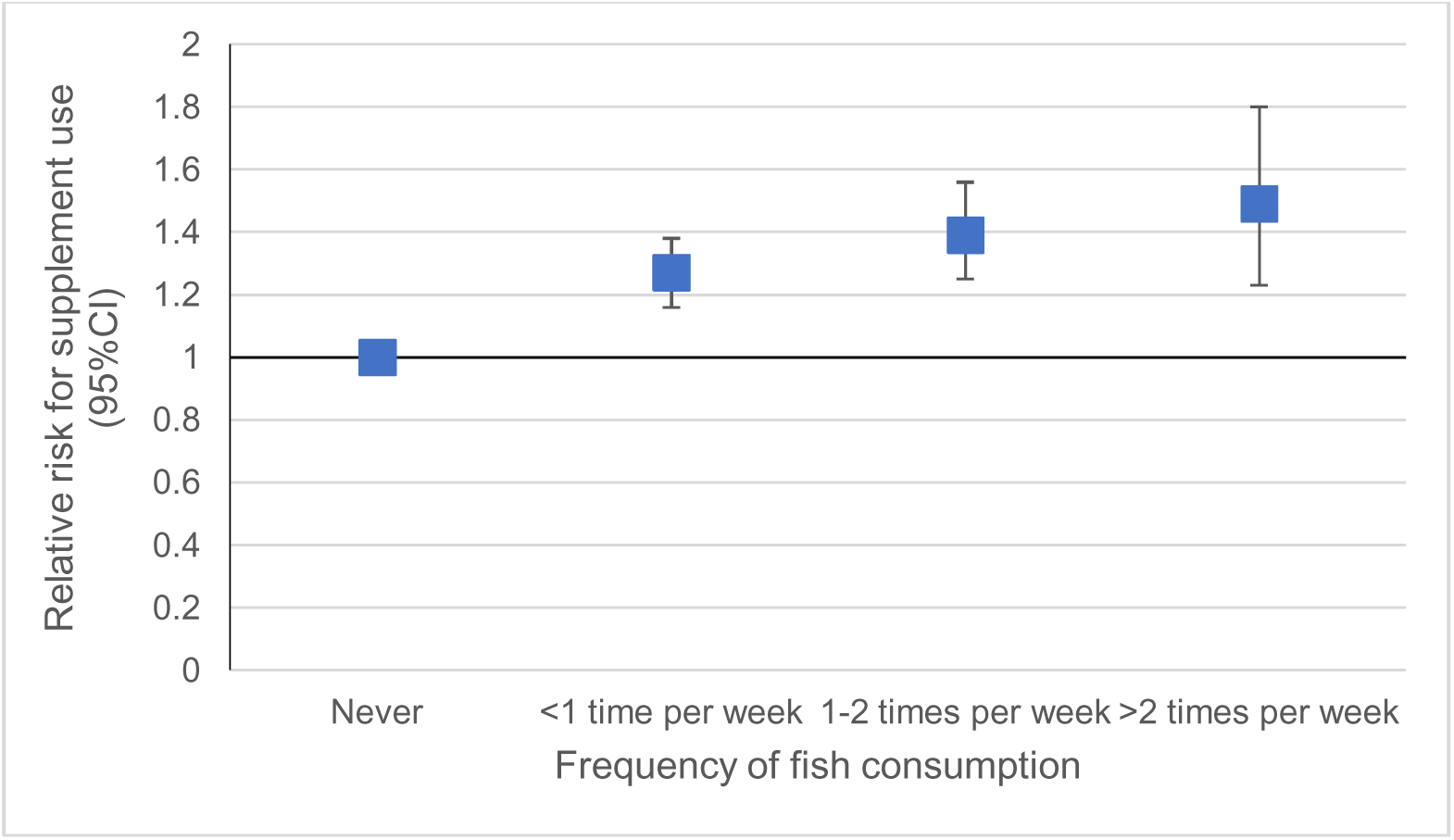
Likelihood of omega-3 polyunsaturated supplement use in pregnancy according to fish consumption during pregnancy within the Environmental influences on Child Health Outcomes (ECHO) cohort.

**Table 2:**
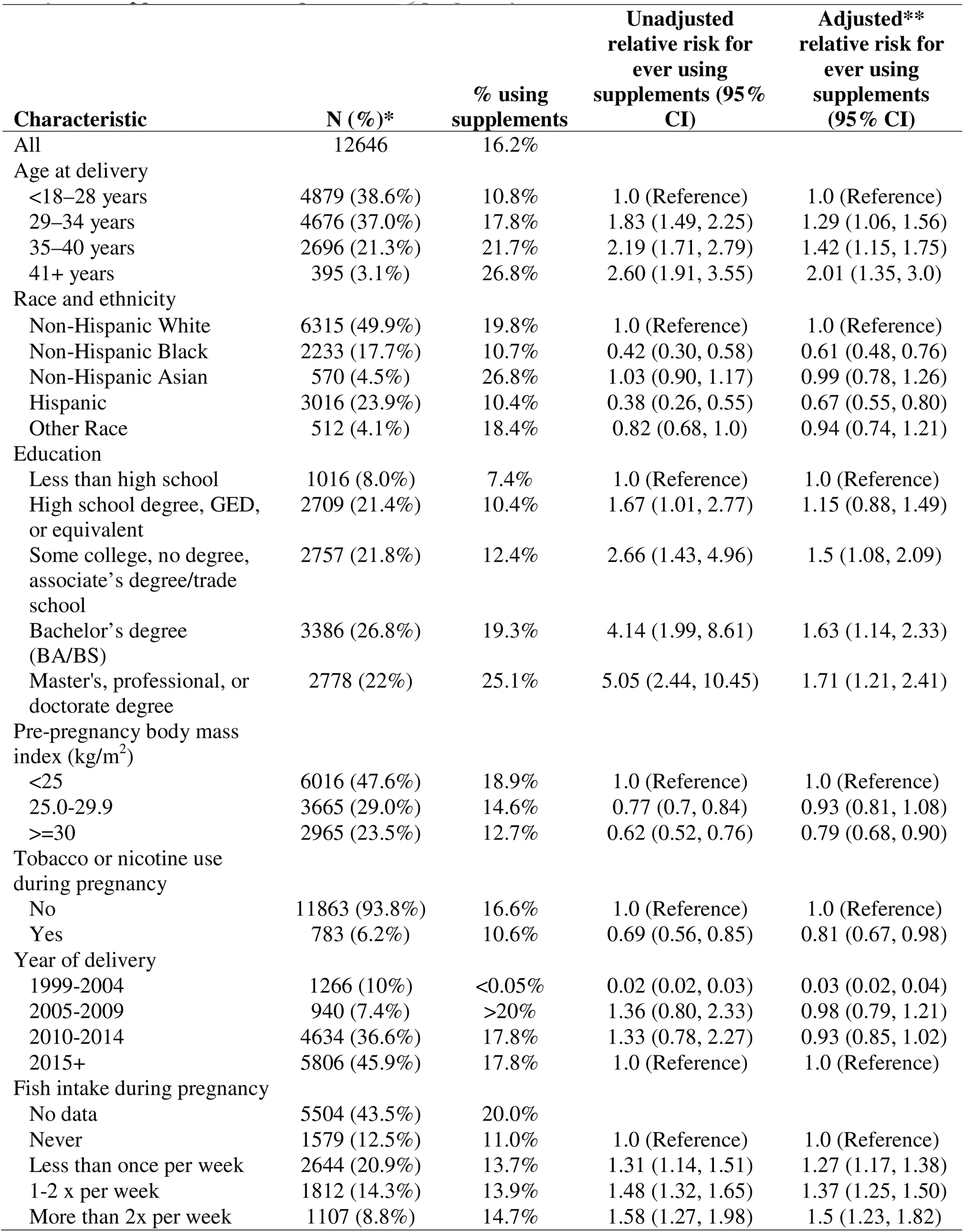
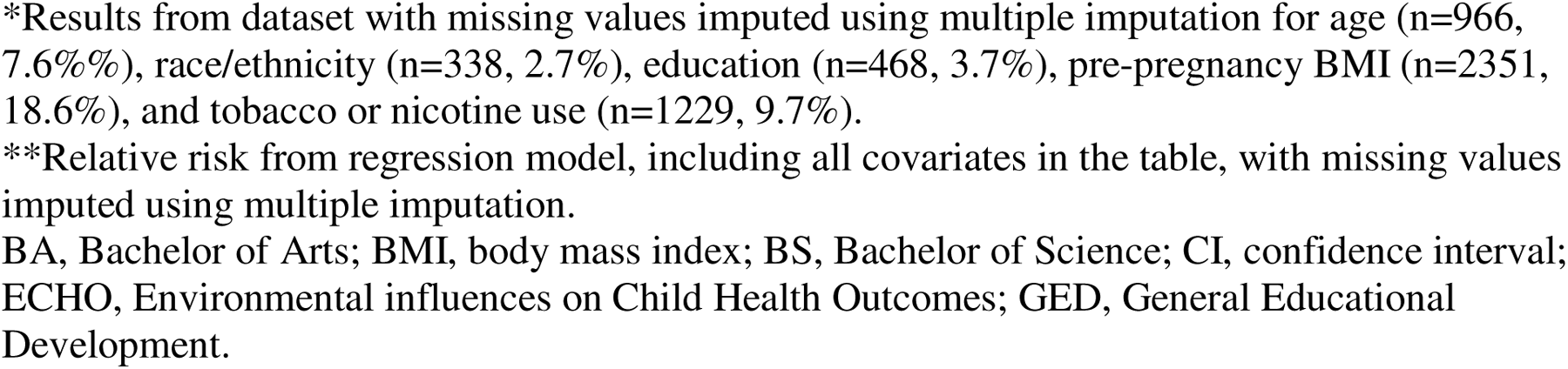
Characteristics of 12,646 ECHO-wide cohort participants with information on omega-3 fatty acid supplement consumption during pregnancy.

| Characteristic | N (%)* | % using supplements | Unadjusted relative risk for ever using supplements (95% CI) | Adjusted** relative risk for ever using supplements (95% CI) |
| --- | --- | --- | --- | --- |
| All | 12646 | 16.2% |  |  |
| Age at delivery |  |  |  |  |
| <18–28 years | 4879 (38.6%) | 10.8% | 1.0 (Reference) | 1.0 (Reference) |
| 29–34 years | 4676 (37.0%) | 17.8% | 1.83 (1.49, 2.25) | 1.29 (1.06, 1.56) |
| 35–40 years | 2696 (21.3%) | 21.7% | 2.19 (1.71, 2.79) | 1.42 (1.15, 1.75) |
| 41+ years | 395 (3.1%) | 26.8% | 2.60 (1.91, 3.55) | 2.01 (1.35, 3.0) |
| Race and ethnicity |  |  |  |  |
| Non-Hispanic White | 6315 (49.9%) | 19.8% | 1.0 (Reference) | 1.0 (Reference) |
| Non-Hispanic Black | 2233 (17.7%) | 10.7% | 0.42 (0.30, 0.58) | 0.61 (0.48, 0.76) |
| Non-Hispanic Asian | 570 (4.5%) | 26.8% | 1.03 (0.90, 1.17) | 0.99 (0.78, 1.26) |
| Hispanic | 3016 (23.9%) | 10.4% | 0.38 (0.26, 0.55) | 0.67 (0.55, 0.80) |
| Other Race | 512 (4.1%) | 18.4% | 0.82 (0.68, 1.0) | 0.94 (0.74, 1.21) |
| Education |  |  |  |  |
| Less than high school | 1016 (8.0%) | 7.4% | 1.0 (Reference) | 1.0 (Reference) |
| High school degree, GED, or equivalent | 2709 (21.4%) | 10.4% | 1.67 (1.01, 2.77) | 1.15 (0.88, 1.49) |
| Some college, no degree, associate's degree/trade school | 2757 (21.8%) | 12.4% | 2.66 (1.43, 4.96) | 1.5 (1.08, 2.09) |
| Bachelor's degree (BA/BS) | 3386 (26.8%) | 19.3% | 4.14 (1.99, 8.61) | 1.63 (1.14, 2.33) |
| Master's, professional, or doctorate degree | 2778 (22%) | 25.1% | 5.05 (2.44, 10.45) | 1.71 (1.21, 2.41) |
| Pre-pregnancy body mass index (kg/m<sup>2</sup>) |  |  |  |  |
| <25 | 6016 (47.6%) | 18.9% | 1.0 (Reference) | 1.0 (Reference) |
| 25.0-29.9 | 3665 (29.0%) | 14.6% | 0.77 (0.7, 0.84) | 0.93 (0.81, 1.08) |
| >=30 | 2965 (23.5%) | 12.7% | 0.62 (0.52, 0.76) | 0.79 (0.68, 0.90) |
| Tobacco or nicotine use during pregnancy |  |  |  |  |
| No | 11863 (93.8%) | 16.6% | 1.0 (Reference) | 1.0 (Reference) |
| Yes | 783 (6.2%) | 10.6% | 0.69 (0.56, 0.85) | 0.81 (0.67, 0.98) |
| Year of delivery |  |  |  |  |
| 1999-2004 | 1266 (10%) | <0.05% | 0.02 (0.02, 0.03) | 0.03 (0.02, 0.04) |
| 2005-2009 | 940 (7.4%) | >20% | 1.36 (0.80, 2.33) | 0.98 (0.79, 1.21) |
| 2010-2014 | 4634 (36.6%) | 17.8% | 1.33 (0.78, 2.27) | 0.93 (0.85, 1.02) |
| 2015+ | 5806 (45.9%) | 17.8% | 1.0 (Reference) | 1.0 (Reference) |
| Fish intake during pregnancy |  |  |  |  |
| No data | 5504 (43.5%) | 20.0% |  |  |
| Never | 1579 (12.5%) | 11.0% | 1.0 (Reference) | 1.0 (Reference) |
| Less than once per week | 2644 (20.9%) | 13.7% | 1.31 (1.14, 1.51) | 1.27 (1.17, 1.38) |
| 1-2 x per week | 1812 (14.3%) | 13.9% | 1.48 (1.32, 1.65) | 1.37 (1.25, 1.50) |
| More than 2x per week | 1107 (8.8%) | 14.7% | 1.58 (1.27, 1.98) | 1.5 (1.23, 1.82) |

\*Results from dataset with missing values imputed using multiple imputation for age (n=966, 7.6% %), race/ethnicity (n=338, 2.7%), education (n=468, 3.7%), pre-pregnancy BMI (n=2351, 18.6%), and tobacco or nicotine use (n=1229, 9.7%).
\*\*Relative risk from regression model, including all covariates in the table, with missing values imputed using multiple imputation.
BA, Bachelor of Arts; BMI, body mass index; BS, Bachelor of Science; CI, confidence interval; ECHO, Environmental influences on Child Health Outcomes; GED, General Educational Development.

## DISCUSSION

A large number of observational studies and some randomized trials have examined associations of total prenatal fish or omega-3 PUFA intake with a range of outcomes.(^21^) Expert opinion has coalesced around the benefits of regular fish or supplement consumption to achieve an intake of at least 500 mg/day of long-chain omega-3 PUFAs (including 200 mg/day of DHA).(^22^) Using data from over 10,000 pregnancies across the U.S. occurring from 1999 through 2020, we observed that almost a quarter of women reported never consuming fish, and only 16% consumed any omega-3 fatty acid supplements. Additionally, fish intake correlated with demographic and health characteristics, albeit somewhat less strongly and not entirely similar to supplement use. Similar to supplement use, fish intake was higher in those who were older and had a higher income and education, but different from supplements, fish intake was higher in those with racial/ethnic identities other than non-Hispanic White and in those who used tobacco and nicotine products. Supplement intake tracked even more strongly with demographics, with the highest likelihood of intake among those who were older, had a higher education and income, and were non-Hispanic White or Asian. Additionally, supplement use was less common among those at higher risk for adverse pregnancy outcomes as a function of using tobacco or nicotine products or having a higher BMI.

Limited recent information is available about fish intake in U.S. pregnancies, yet such estimates are essential for efforts to model health risks and benefits from nutrients and contaminants commonly found in fish. For example, in the most recent FDA assessment of the net effects of eating commercial fish on fetal neurodevelopment, which was conducted in 2014, the FDA estimated the types and amount of fish that people eat.(^5^) This estimation was based on three sources of data that were collected ∼15, ∼20, and ∼30 years ago: National Marine Fisheries Service market share data on consumable commercial fish from 2007, NHANES data from 1999-2000, and U.S. Department of Agriculture’s Continuing Survey of Food Intake by Individuals (CSFII) data from 1989-1991. With these data, the FDA estimated that women of childbearing age in the U.S. consumed a mean of 3.7 and a median of 1.9 ounces of fish per week. It is notable that this model was based entirely on dietary data from non-pregnant persons and assumed that pregnant women eat a similar amount, despite evidence that women consume less fish during pregnancy.(^6,23^) Further, fish consumption in women of childbearing age may have decreased since these data were collected.(^8,9^)

Evidence to support routine omega-3 supplementation in pregnancy has not been entirely consistent.(^1,24^) Nevertheless, those with low baseline fish intake or omega-3 PUFA status,(^25,26^) or a BMI that indicates obesity(^27,28^) may particularly benefit from supplementation. We did not observe that supplement use was more common in those with low fish intake or a high BMI, but rather the opposite. In another study of ECHO participants, more than 99% of the sample reported use of vitamins and minerals containing supplements during pregnancy, but that analysis did not include omega-3 PUFA supplements.(^29^) In contrast to vitamin and mineral supplement use, which abated most nutrient risk disparities from diet alone, we found that omega-3 PUFA supplement use was less common among those who did not eat fish.

The large sample size is a strength, and we included data from over the past two decades up to 2020. In contrast, published NHANES analyses include about 1,000 pregnancies and a decade of data extending to 2015. Limitations include our inability to assess specific fish types, given the varied dietary assessment instruments used across cohorts, or to assign intake by trimester. However, most prior studies have examined total fish intake, and current US guidelines recommend total fish intake rather than specific subtypes such as “fatty” fish.^2^ Also, although different studies administered different questionnaires, we accounted for the number of questions asked about fish. Additionally, we do not have information on supplement dose. Both fish and supplement intake were self-reported, as is typical in studies of usual diet, and reporting may have been biased.

The ECHO population is nationwide but not necessarily nationally representative,^19^ as it draws upon individuals who elected to enroll in cohorts and who may be more health conscious than the general population. The very low fish and omega-3 PUFA supplement intake we observed may overestimate actual use in all U.S. pregnancies, as more health-conscious persons may consume more fish and supplements, or alternatively, it may be that more health-conscious persons try to avoid mercury exposure from fish. Our results are especially timely given that both the World Health Organization and US National Academies are currently evaluating the evidence on fish intake in pregnancy.^30,31^ Ongoing effective public health advice and resources to support clinicians,^32,33^ are needed to encourage consumption of low-mercury fish during pregnancy and intake of omega-3 supplements among those who do not consume fish.

## Data Availability

De-identified ECHO-wide cohort data are publicly available via the NICHD Data and Specimen Hub (DASH).

https://dash.nichd.nih.gov/study/417122

## CRediT Author Statement

*Conceptualization*: Oken, Lyall, Musci, Westlake

*Methodology*: Oken, Lyall, Musci, Westlake, Gachigi

*Software*: Westlake, Gachigi

*Validation*: Musci, Westlake, Gachigi

*Formal analysis*: Musci, Westlake, Gachigi

*Investigation*: Oken, Aschner, Barnes, Bastain, Buss, Camargo, Cordero, Dabelea, Dunlop, Ghassabian, Hipwell, Hockett, Karagas, Lugo-Candelas, Margolis, O’Connor, Shuster, Straughen, Lyall

*Resources*: Oken, Aschner, Barnes, Bastain, Buss, Camargo, Cordero, Dabelea, Dunlop, Ghassabian, Hipwell, Hockett, Karagas, Lugo-Candelas, Margolis, O’Connor, Shuster, Straughen, Lyall

*Data Curation*: Westlake

*Writing, Original Draft*: Oken

*Writing, Review and Editing*: All

*Visualization*: Oken

*Supervision*: Oken, Aschner, Barnes, Bastain, Buss, Camargo, Cordero, Dabelea, Dunlop, Ghassabian, Hipwell, Hockett, Karagas, Lugo-Candelas, Margolis, O’Connor, Shuster, Straughen, Lyall

*Project Administration*: Oken and Lyall

*Funding Acquisition*: Oken, Aschner, Barnes, Bastain, Buss, Camargo, Cordero, Dabelea, Dunlop, Ghassabian, Hipwell, Hockett, Karagas, Lugo-Candelas, Margolis, O’Connor, Shuster, Straughen, Lyall

## ACKNOWLEDGMENTS

The authors would like to acknowledge the unique contributions of Zhumin Zhang, MS, PhD, University of Wisconsin-Madison, to this study, which included investigation, resources, supervision, and funding acquisition.

The authors wish to thank our ECHO colleagues; the medical, nursing, and program staff; and the children and families participating in the ECHO cohorts. We also acknowledge the contribution of the following ECHO program collaborators:

**ECHO Components**—Coordinating Center: Duke Clinical Research Institute, Durham, North Carolina: Smith PB, Newby KL; Data Analysis Center: Johns Hopkins University Bloomberg School of Public Health, Baltimore, Maryland: Jacobson LP; Research Triangle Institute, Durham, North Carolina: Catellier DJ; Person-Reported Outcomes Core: Northwestern University, Evanston, Illinois: Gershon R, Cella D.

**ECHO Awardees and Cohorts**—Northeastern University, Boston, Massachusetts: Alshawabkeh, AN; Icahn School of Medicine at Mount Sinai, New York, NY: Teitelbaum SL; Stroustrup A; Cohen Children’s Medical Center, Northwell Health: Stroustrup A; Memorial Hospital of Rhode Island, Providence RI: Koinis Mitchell D, Deoni S; New York State Psychiatric Institute, New York, NY: Duarte C; University of Puerto Rico, San Juan, PR: Canino G; Avera Health Rapid City, Rapid City, SD: Elliott A; Kaiser Permanente Northern California Division of Research, Oakland, CA: Ferrara A; University of Wisconsin, Madison WI: Gern J; Henry Ford Health System:, Detroit, MI: Zoratti E; Washington University in St Louis, St Louis, MO: Rivera-Spoljaric K; University of Wisconsin, Madison, WI: Singh A; Vanderbilt University, Nashville, TN: Hartert T; University of Southern California, Los Angeles, CA: Breton C; Farzan S; Habre R; University of California Davis Mind Institute, Sacramento, CA: Hertz-Picciotto I; University of Washington, Department of Environmental and Occupational Health Sciences, Seattle, WA: Karr C; University of Tennessee Health Science Center, Memphis, TN: Mason A; Brigham and Women’s Hospital, Boston, MA: Weiss S; Boston University Medical Center, Boston, MA: O’Connor G; Kaiser Permanente, Southern California, San Diego, CA: Zeiger R; Washington University of St. Louis, St Louis, MO: Bacharier L; University of California, UC Davis Medical Center Mind Institute, Davis, CA: Schmidt R; Johns Hopkins Bloomberg School of Public Health: Volk H; Kaiser Permanente Northern California Division of Research: Croen L; Columbia University Medical Center, New York, NY: Herbstman J; University of Illinois, Beckman Institute, Urbana, IL: Schantz S; University of California, San Francisco:, San Francisco, CA: Woodruff T; New York School of Medicine, New York, NY: Trasande L; Icahn School of Medicine at Mount Sinai, New York, NY: Wright R; Boston Children’s Hospital, Boston MA: Bosquet-Enlow M.

## Disclaimer

The content is solely the responsibility of the authors and does not necessarily represent the official views of the National Institutes of Health.

## Ethical Standards Disclosure

This study was conducted according to the guidelines laid down in the Declaration of Helsinki, and participant activities were overseen and approved by both central and site-specific Institutional Review Boards. All participants provided written informed consent.

