## Supplemental Fig and Supplemental Table 1 for "Demographic and health characteristics associated with fish and n-3 fatty acid supplement intake during pregnancy: results from pregnancy cohorts in the ECHO program"

**SUPPLEMENTARY MATERIAL**

Supplementary Figure 1: Flow of participants in the analysis

Supplementary Table 1: Characteristics of 10,800 ECHO-wide cohort participants with information on fish consumption during pregnancy

Supplementary Figure 1: Flow of participants in the analysis

ECHO pregnancies

N= 56,797 pairs;

N= 69 cohorts

ECHO singleton pregnancies,

1 per pregnant participant	N= 52,333 pairs; N= 69 cohorts

EXCLUDE: multiple births and repeat pregnancies

ECHO singleton pregnancies and prenatal fish data

N= 10,800 pregnancies

N= 23 cohorts

EXCLUDE: no prenatal fish data

EXCLUDE: no prenatal supplement data

ECHO singleton pregnancies and prenatal omega-3 supplement data

N= 12,646 pregnancies

N= 35 cohorts

Supplementary Table 1: Characteristics of 10,800 ECHO-wide cohort participants with information on fish consumption during pregnancy, overall and according to 4 categories of fish consumption during pregnancy

| **Characteristic** | **All participants** | | **According to category of fish consumption during pregnancy** | | | | | | | |
| --- | --- | --- | --- | --- | --- | --- | --- | --- | --- | --- |
|  |  |  | **Never or less than 1 serving per month** | | **Less than 1 serving per week** | | **1-2 servings per week** | | **More than 2 servings per week** | |
|  | **N** | **Column %** | **N** | **Row %** | **N** | **Row %** | **N** | **Row %** | **N** | **Row %** |
| All | 10800 | 100.0 | 2655 | 24.6 | 4335 | 40.1 | 2387 | 22.1 | 1423 | 13.2 |
| Age at delivery |  |  |  |  |  |  |  |  |  |  |
| Missing | 1049 | 9.7 | 354 | 33.7 | 514 | 49.0 | 125 | 11.9 | 56 | 5.3 |
| <18–28 years | 3494 | 324 | 975 | 27.9 | 1363 | 39.0 | 710 | 20.3 | 446 | 12.8 |
| 29–34 years | 3878 | 35.9 | 923 | 23.8 | 1539 | 39.7 | 886 | 22.8 | 530 | 13.7 |
| 35–40 years | 2100 | 19.4 | 359 | 17.1 | 812 | 38.7 | 590 | 28.1 | 339 | 16.1 |
| 41+ years | 279 | 2.6 | 44 | 15.8 | 107 | 38.4 | 76 | 27.2 | 52 | 18.6 |
| Race/ethnicity |  |  |  |  |  |  |  |  |  |  |
| Missing | 586 | 5.4 | 192 | 32.8 | 257 | 43.9 | 89 | 15.2 | 48 | 8.2 |
| Non-Hispanic White | 5055 | 46.8 | 1373 | 27.2 | 2058 | 40.7 | 1101 | 21.8 | 523 | 10.3 |
| Non-Hispanic Black | 1865 | 17.3 | 286 | 15.3 | 732 | 39.2 | 491 | 26.3 | 356 | 19.1 |
| Non-Hispanic Asian | 605 | 5.6 | 100 | 16.5 | 166 | 27.4 | 180 | 29.8 | 159 | 26.3 |
| Hispanic | 2166 | 20.1 | 565 | 26.1 | 913 | 42.2 | 415 | 19.2 | 273 | 12.6 |
| Other Race | 523 | 4.8 | 139 | 26.6 | 209 | 40.0 | 111 | 21.2 | 64 | 12.2 |
| Annual household income |  |  |  |  |  |  |  |  |  |  |
| Missing | 5513 | 51.1 | 1249 | 22.7 | 2165 | 39.3 | 1334 | 24.2 | 765 | 13.9 |
| <$30,000 | 1801 | 16.7 | 454 | 25.2 | 713 | 39.6 | 362 | 20.1 | 272 | 15.1 |
| $30,000-$49,999 | 668 | 6.2 | 188 | 28.1 | 284 | 42.5 | 133 | 19.9 | 63 | 9.4 |
| $50,000-$74,999 | 758 | 7.0 | 216 | 28.5 | 306 | 40.4 | 145 | 19.1 | 91 | 12.0 |
| $75,000-$99,999 | 605 | 5.6 | 200 | 33.1 | 247 | 40.8 | 110 | 18.2 | 48 | 7.9 |
| $100,000 or more | 1455 | 13.5 | 348 | 23.9 | 620 | 42.6 | 303 | 20.8 | 184 | 12.6 |

|  | **All participants** | | **According to category of fish consumption during pregnancy** | | | | | | | |
| --- | --- | --- | --- | --- | --- | --- | --- | --- | --- | --- |
|  |  |  | **Never or less than 1 serving per month** | | **Less than 1 serving per week** | | **1-2 servings per week** | | **More than 2 servings per week** | |
| **Characteristic** | **N** | **Column %** | **N** | **Row %** | **N** | ***P*-value** | **N** | **Row %** | **N** | **Row %** |
| Education |  |  |  |  |  |  |  |  |  |  |
| Missing | 1048 | 9.7 | 329 | 31.4 | 460 | 43.9 | 174 | 16.6 | 85 | 8.1 |
| Less than high school | 597 | 5.5 | 142 | 23.8 | 240 | 40.2 | 120 | 20.1 | 95 | 15.9 |
| High school degree, GED, or equivalent | 1845 | 17.1 | 456 | 24.7 | 761 | 41.2 | 376 | 20.4 | 252 | 13.7 |
| Some college, no degree, associate’s degree/trade school | 2408 | 22.3 | 643 | 26.7 | 942 | 39.1 | 498 | 20.7 | 325 | 13.5 |
| Bachelor’s degree (BA, BS) | 2848 | 26.4 | 649 | 22.8 | 1127 | 39.6 | 687 | 24.1 | 385 | 13.5 |
| Master's, professional, or doctorate degree | 2054 | 19.0 | 436 | 21.2 | 805 | 39.2 | 532 | 25.9 | 281 | 13.7 |
| Pre-pregnancy body mass index (kg/m2) |  |  |  |  |  |  |  |  |  |  |
| Missing | 2525 | 23.4 | 795 | 31.5 | 1108 | 43.9 | 414 | 16.4 | 208 | 8.2 |
| <18.5 | 242 | 2.2 | 66 | 27.3 | 92 | 38.0 | 52 | 21.5 | 32 | 13.2 |
| 18.5-24.9 | 3742 | 34.7 | 776 | 20.7 | 1442 | 38.5 | 956 | 25.5 | 568 | 15.2 |
| 25-29.9 | 2117 | 19.6 | 505 | 23.9 | 825 | 39.0 | 488 | 23.1 | 299 | 14.1 |
| ≥30 | 2174 | 20.1 | 513 | 23.6 | 868 | 39.9 | 477 | 21.9 | 316 | 14.5 |
| Prenatal tobacco or nicotine use |  |  |  |  |  |  |  |  |  |  |
| Missing | 2650 | 24.5 | 943 | 35.6 | 1245 | 47.0 | 329 | 12.4 | 133 | 5.0 |
| No | 7694 | 71.2 | 1626 | 21.1 | 2924 | 38.0 | 1942 | 25.2 | 1202 | 15.6 |
| Yes | 456 | 4.2 | 86 | 18.9 | 166 | 36.4 | 116 | 25.4 | 88 | 19.3 |
| Year of delivery |  |  |  |  |  |  |  |  |  |  |
| 1999-2004 | 1161 | 10.8 | 139 | 12.0 | 430 | 37.0 | 375 | 32.2 | 217 | 18.7 |
| 2005-2009 | 942 | 8.7 | 157 | 16.7 | 356 | 37.8 | 261 | 27.7 | 168 | 17.8 |
| 2010-2014 | 3164 | 29.3 | 616 | 19.5 | 1094 | 34.6 | 872 | 27.6 | 582 | 18.4 |
| 2015+ | 5533 | 51.2 | 1743 | 31.5 | 2455 | 44.4 | 879 | 15.9 | 456 | 8.2 |
| Number of fish questions on questionnaire |  |  |  |  |  |  |  |  |  |  |
| 1 | 3712 | 34.4 | 1396 | 37.6 | 1757 | 47.3 | 421 | 11.3 | 138 | 3.7 |
| More than 1 | 7088 | 65.6 | 1259 | 17.8 | 2578 | 36.4 | 1966 | 27.7 | 1285 | 18.1 |

BA, Bachelor of Arts; BMI, body mass index; BS, Bachelor of Science; GED, General Educational Development.
